# Healthcare workers’ views on mandatory SARS-CoV-2 vaccination in the United Kingdom: findings from the UK-REACH prospective longitudinal cohort study

**DOI:** 10.1101/2022.01.11.22269017

**Authors:** Katherine Woolf, Mayuri Gogoi, Christopher A Martin, Padmasayee Papineni, Susie Lagrata, Laura B Nellums, I Chris McManus, Anna L Guyatt, Carl Melbourne, Luke Bryant, Amit Gupta, Catherine John, Sue Carr, Martin D Tobin, Sandra Simpson, Bindu Gregary, Avinash Aujayeb, Stephen Zingwe, Rubina Reza, Laura J Gray, Kamlesh Khunti, Manish Pareek, On behalf of the UK-REACH Study Collaborative Group

## Abstract

**Background:** Several countries now have mandatory SARS-CoV-2/COVID-19 vaccination for healthcare workers (HCWs) or the general population. HCWs’ views on this are largely unknown.

**Methods:** We administered an online questionnaire to 17891 United Kingdom (UK) HCWs in Spring 2021 as part of the United Kingdom Research study into Ethnicity And COVID-19 outcomes in Healthcare workers (UK-REACH) nationwide prospective cohort study. We categorised responses to a free-text question “What should society do if people don’t get vaccinated against COVID-19?” using content analysis. We collapsed categories into a binary variable: favours mandatory vaccination or not and used logistic regression to calculate its demographic predictors, and occupational, health and attitudinal predictors adjusted for demographics.

**Findings:** Of 5633 questionnaire respondents, 3235 answered the freetext question; 18% (n=578) of those favoured mandatory vaccination but the most frequent suggestion was education (32%, n=1047). Older HCWs, HCWs vaccinated against influenza (OR 1.48; 95%CI 1.10 – 1.99, vs none) and with more positive vaccination attitudes generally (OR 1.10; 95%CI 1.06 – 1.14) were more likely to favour mandatory vaccination (OR 1.26; 95%CI 1.17 – 1.37, per decade increase), whereas female HCWs (OR= 0.80, 95%CI 0.65 – 0.99, vs male), Black HCWs (OR= 0.48, 95%CI 0.26 – 0.87, vs White), those hesitant about COVID-19 vaccination (OR= 0.56; 95%CI 0.43 – 0.71, vs not hesitant), in an Allied Health Profession (OR 0.67; 95%CI 0.51 – 0.88, vs Medical), or who trusted their organisation (OR 0.78; 95%CI 0.63 – 0.96) were less likely to.

**Interpretation:** Only one in six of the HCWs in this large, diverse, UK-wide sample favoured mandatory vaccination. Building trust, educating and supporting HCWs who are hesitant about vaccination may be more acceptable, effective and equitable.

**Funding:** MRC-UK Research and Innovation grant (MR/V027549/1) and the Department of Health and Social Care via the National Institute for Health Research.

## Introduction

Vaccines against Coronavirus disease 2019 (COVID-19) were approved for emergency use in the United Kingdom (UK) in December 2020 and since then have been offered in a staggered manner to everyone aged 12 years and over.^1^ Figures released by the UK government show that by 6^th^ January 2022, 92% of adults in England had their first dose of a COVID-19 vaccine, 88% had had a second dose and 68% had had a booster or third dose.^2^ While these figures look promising, they mask considerable variations in vaccination uptake by region and demographics,^3^ with vaccine uptake lower in certain ethnic groups, among women, and in younger groups.^4,5^ This patterning of vaccine uptake is also observed among HCWs. ^2,6^

With COVID-19 cases surging and the emergence of the Omicron variant of concern, Italy, Greece and France have made COVID-19 vaccine compulsory for healthcare workers (HCWs),^7,8^ and Austria and Greece have made vaccinations mandatory for the general population.^9,10^ The United States has mandated COVID-19 vaccination for all federal employees including healthcare personnel.^11^ In England, COVID-19 vaccination was made mandatory for social care workers in November 2021, and from April 2022 it will also become mandatory for HCWs working in other healthcare settings.^12,13^

The introduction of strict measures to improve vaccine uptake has given rise to a variety of views, with some agreeing these measures are for ‘greater good’,^14^ while others fear they will deepen vaccine hesitancy and mistrust.^15^ Opinion polls from the UK demonstrate that the majority of the population agrees with mandatory COVID-19 vaccination for the general public and for National Health Service (NHS) or social care staff.^16,17^ By contrast, the World Health Organisation (WHO) has cautioned against mandatory vaccination, with the Director of WHO Europe, Hans Kluge, stating that, “mandates should never contribute to increasing social inequalities in access to health and social services”.^18^ Several HCW regulators and representative bodies in the UK have stated concerns about mandatory vaccination for HCWs, including that it could erode trust and exacerbate existing workforce shortages.^19-22^ The General Medical Council, UK have also identified that potential issues could arise among vaccinators if they feel that an individual’s choice to receive a vaccination is unduly influenced by a deployment requirement.^23^

Previous research has demonstrated mixed views among HCWs regarding mandatory vaccinations against other diseases, particularly influenza, and support for mandatory vaccination varies by country, occupational group and vaccination status.^24^’^25^ At present there is little research on HCW views on compulsory COVID-19 vaccination. Understanding HCWs views on this topic (as well as on other potential ways of increasing COVID-19 vaccine uptake such as vaccine passports) is important as HCWs have considerable influence on the public’s intention to get vaccinated^26^ and they also deliver vaccinations. Capturing HCWs’ suggestions for how to improve vaccine uptake could also help address vaccine hesitancy, shape policy and generate support for policy measures. In this study, we aimed to examine the views of HCWs in the UK about mandatory vaccination against COVID-19 for the UK population and/or for health and social care workers specifically, collected as part of the United Kingdom Research study into Ethnicity And COVID-19 outcomes among Healthcare workers (UK-REACH).

## Methods

### Overview

UK-REACH is a programme of work aiming to determine the impact of the COVID-19 pandemic on UK HCWs, and establish whether this differs according to ethnicity. This analysis uses data from the baseline questionnaire of the prospective nationwide cohort study, administered between December 2020 and March 2021 and the first follow up questionnaire, administered between 21^st^ April 2021 and 26^th^ June 2021. See study protocol^27^ for details.

### Study population

We recruited individuals aged 16 years or over, living in the UK and employed as clinical or ancillary workers in a healthcare setting and/or registered with one of seven UK professional regulatory bodies. Recruitment into the study has been described extensively in previous work.^6,27,28^

This study used data from the second UK-REACH questionnaire, which was administered online to the 17,891 people who responded to the first questionnaire. HCWs were included in the analysed cohort if they answered both the baseline and the first follow up questionnaire.

### Qualitative methods

In the follow-up questionnaire, participants could respond to a free text question: “What should society do if people don’t get vaccinated against COVID-19?” We used the qualitative method of manifest content analysis,^29,30^ using an inductive (data-driven) approach to develop and assign codes to each response.

One researcher (KW) read through half of the responses and identified codes arising from the data, refining them iteratively throughout this process. When a participant’s response included more than one code, we used the most socially restrictive code, for example, if a participant said that people should be educated and also that HCWs should have mandatory vaccinations, we coded this as mandatory vaccination for HCWs. This resulted in a dataset in which each participant’s response had a single code. A second researcher (MG) then independently coded the remaining responses using the same coding framework, and double-coded 362 (25%) of KW’s coded responses blindly. We assessed consistency and inter-coder reliability using the following formula^31^:

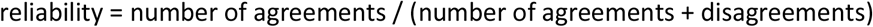

Thereafter, both researchers discussed the coding framework, their interpretations of the codes, and the disagreements and finally arrived at an agreed code for each response.

### Statistical methods

#### Outcome measures

We used two outcome measures for the quantitative analysis. The first was a multinomial categorical variable with one level for each code (see Table 1). We collapsed this into a binary outcome variable ‘favours mandatory vaccination’ (1=favours mandatory vaccination for the general public or for HCWs, 0=all other codes).

**Table 1.**
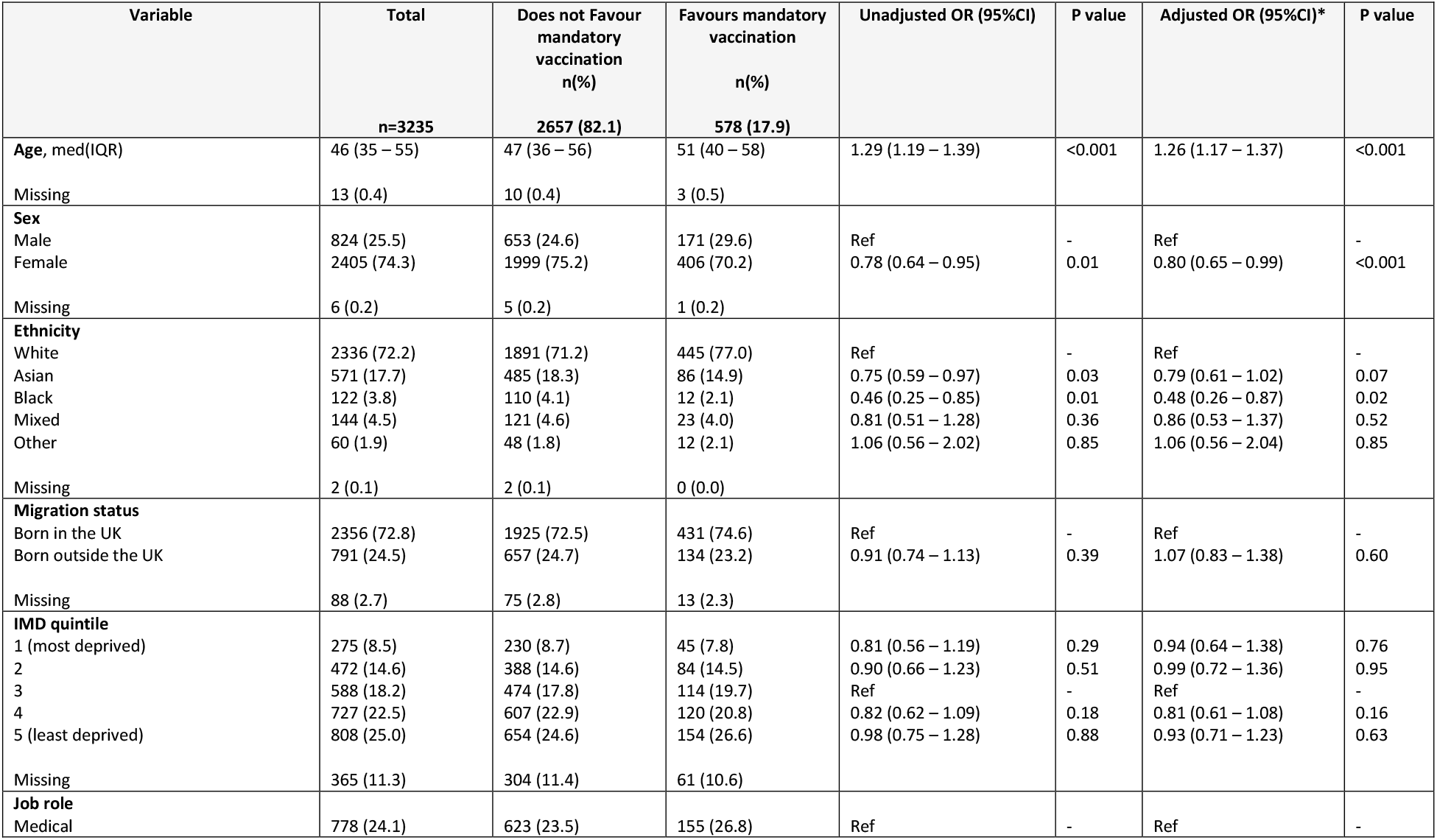
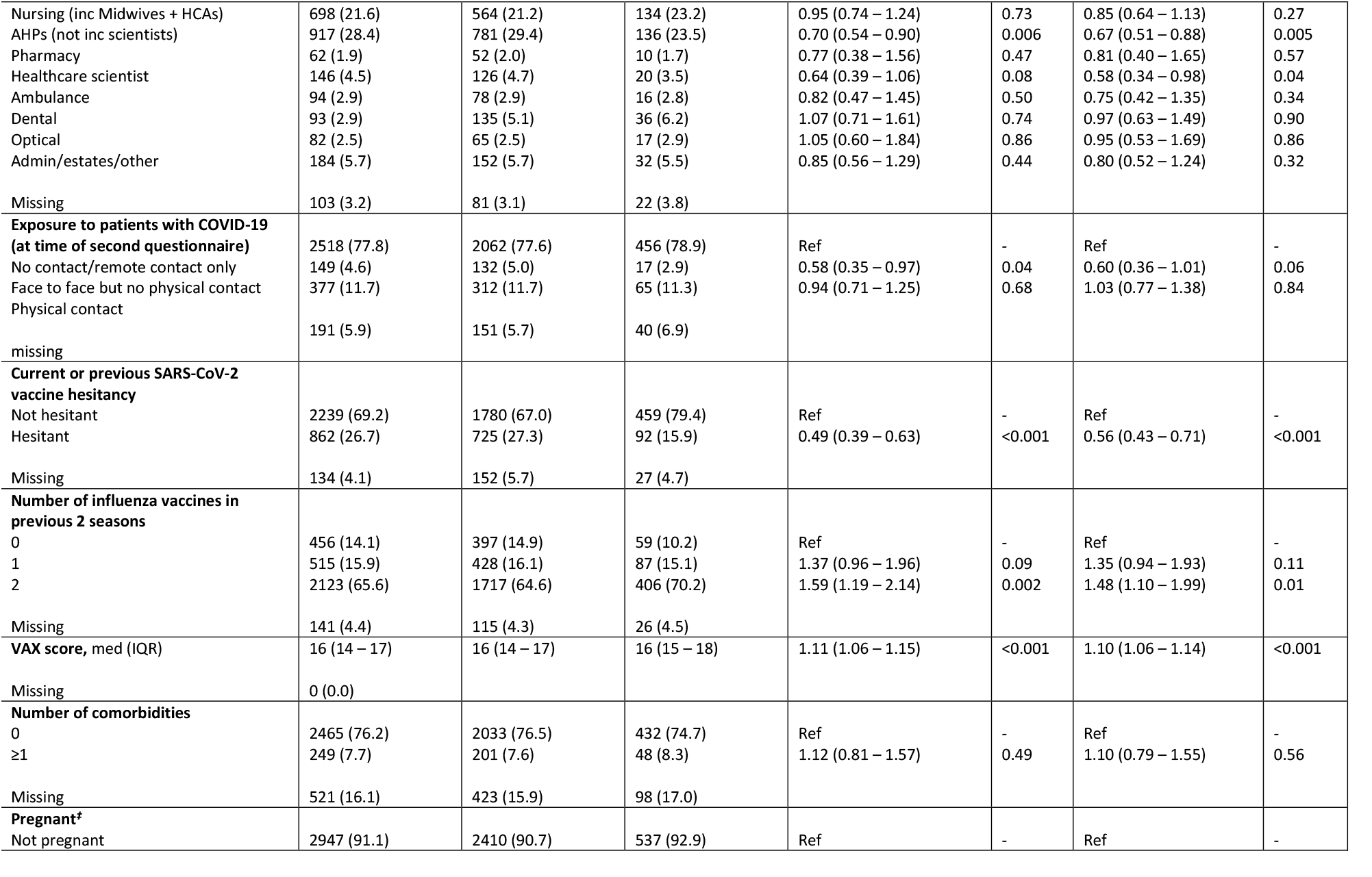
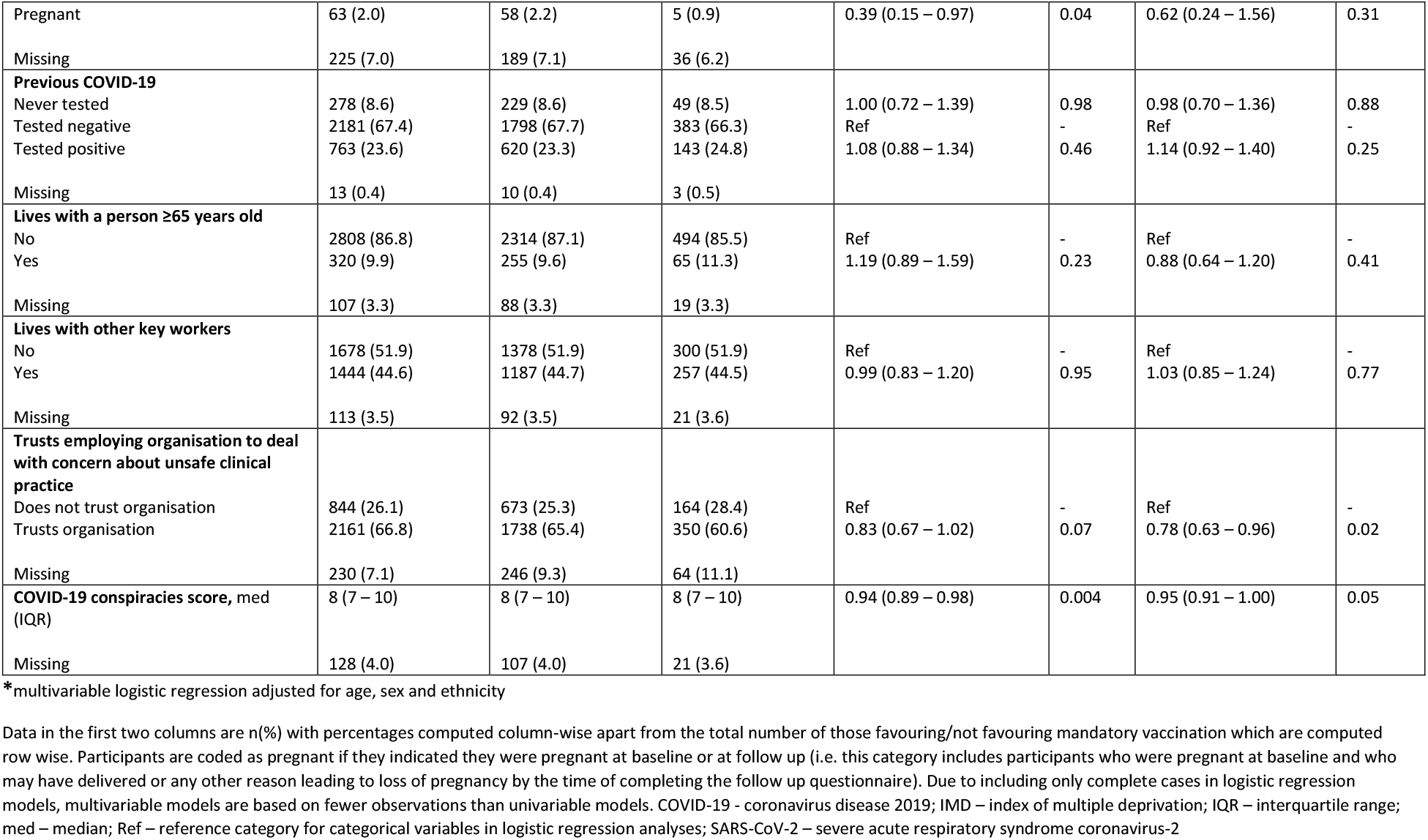
Description of the cohort stratified by the binary outcome of favouring mandatory vaccination or not and selected predictor variables, together with unadjusted and adjusted odds ratios for the association of predictor variables with the outcome.

| Variable | Total<br>n=3235 | Does not Favour<br>mandatory<br>vaccination<br>n(%)<br>2657 (82.1) | Favours mandatory<br>vaccination<br>n(%)<br>578 (17.9) | Unadjusted OR (95%CI) | P value | Adjusted OR (95%CI)* | P value |
| --- | --- | --- | --- | --- | --- | --- | --- |
| **Age, med(IQR)** | 46 (35 – 55) | 47 (36 – 56) | 51 (40 – 58) | 1.29 (1.19 – 1.39) | <0.001 | 1.26 (1.17 – 1.37) | <0.001 |
| Missing | 13 (0.4) | 10 (0.4) | 3 (0.5) |  |  |  |  |
| **Sex** |  |  |  |  |  |  |  |
| Male | 824 (25.5) | 653 (24.6) | 171 (29.6) | Ref | - | Ref | - |
| Female | 2405 (74.3) | 1999 (75.2) | 406 (70.2) | 0.78 (0.64 – 0.95) | 0.01 | 0.80 (0.65 – 0.99) | <0.001 |
| Missing | 6 (0.2) | 5 (0.2) | 1 (0.2) |  |  |  |  |
| **Ethnicity** |  |  |  |  |  |  |  |
| White | 2336 (72.2) | 1891 (71.2) | 445 (77.0) | Ref | - | Ref | - |
| Asian | 571 (17.7) | 485 (18.3) | 86 (14.9) | 0.75 (0.59 – 0.97) | 0.03 | 0.79 (0.61 – 1.02) | 0.07 |
| Black | 122 (3.8) | 110 (4.1) | 12 (2.1) | 0.46 (0.25 – 0.85) | 0.01 | 0.48 (0.26 – 0.87) | 0.02 |
| Mixed | 144 (4.5) | 121 (4.6) | 23 (4.0) | 0.81 (0.51 – 1.28) | 0.36 | 0.86 (0.53 – 1.37) | 0.52 |
| Other | 60 (1.9) | 48 (1.8) | 12 (2.1) | 1.06 (0.56 – 2.02) | 0.85 | 1.06 (0.56 – 2.04) | 0.85 |
| Missing | 2 (0.1) | 2 (0.1) | 0 (0.0) |  |  |  |  |
| **Migration status** |  |  |  |  |  |  |  |
| Born in the UK | 2356 (72.8) | 1925 (72.5) | 431 (74.6) | Ref | - | Ref | - |
| Born outside the UK | 791 (24.5) | 657 (24.7) | 134 (23.2) | 0.91 (0.74 – 1.13) | 0.39 | 1.07 (0.83 – 1.38) | 0.60 |
| Missing | 88 (2.7) | 75 (2.8) | 13 (2.3) |  |  |  |  |
| **IMD quintile** |  |  |  |  |  |  |  |
| 1 (most deprived) | 275 (8.5) | 230 (8.7) | 45 (7.8) | 0.81 (0.56 – 1.19) | 0.29 | 0.94 (0.64 – 1.38) | 0.76 |
| 2 | 472 (14.6) | 388 (14.6) | 84 (14.5) | 0.90 (0.66 – 1.23) | 0.51 | 0.99 (0.72 – 1.36) | 0.95 |
| 3 | 588 (18.2) | 474 (17.8) | 114 (19.7) | Ref | - | Ref | - |
| 4 | 727 (22.5) | 607 (22.9) | 120 (20.8) | 0.82 (0.62 – 1.09) | 0.18 | 0.81 (0.61 – 1.08) | 0.16 |
| 5 (least deprived) | 808 (25.0) | 654 (24.6) | 154 (26.6) | 0.98 (0.75 – 1.28) | 0.88 | 0.93 (0.71 – 1.23) | 0.63 |
| Missing | 365 (11.3) | 304 (11.4) | 61 (10.6) |  |  |  |  |
| **Job role** |  |  |  |  |  |  |  |
| Medical | 778 (24.1) | 623 (23.5) | 155 (26.8) | Ref | - | Ref | - |
| Nursing (inc Midwives + HCAs) | 698 (21.6) | 564 (21.2) | 134 (23.2) | 0.95 (0.74 – 1.24) | 0.73 | 0.85 (0.64 – 1.13) | 0.27 |
| AHPs (not inc scientists) | 917 (28.4) | 781 (29.4) | 136 (23.5) | 0.70 (0.54 – 0.90) | 0.006 | 0.67 (0.51 – 0.88) | 0.005 |
| Pharmacy | 62 (1.9) | 52 (2.0) | 10 (1.7) | 0.77 (0.38 – 1.56) | 0.47 | 0.81 (0.40 – 1.65) | 0.57 |
| Healthcare scientist | 146 (4.5) | 126 (4.7) | 20 (3.5) | 0.64 (0.39 – 1.06) | 0.08 | 0.58 (0.34 – 0.98) | 0.04 |
| Ambulance | 94 (2.9) | 78 (2.9) | 16 (2.8) | 0.82 (0.47 – 1.45) | 0.50 | 0.75 (0.42 – 1.35) | 0.34 |
| Dental | 93 (2.9) | 135 (5.1) | 36 (6.2) | 1.07 (0.71 – 1.61) | 0.74 | 0.97 (0.63 – 1.49) | 0.90 |
| Optical | 82 (2.5) | 65 (2.5) | 17 (2.9) | 1.05 (0.60 – 1.84) | 0.86 | 0.95 (0.53 – 1.69) | 0.86 |
| Admin/estates/other | 184 (5.7) | 152 (5.7) | 32 (5.5) | 0.85 (0.56 – 1.29) | 0.44 | 0.80 (0.52 – 1.24) | 0.32 |
| Missing | 103 (3.2) | 81 (3.1) | 22 (3.8) |  |  |  |  |
| **Exposure to patients with COVID-19 (at time of second questionnaire)** |  |  |  |  |  |  |  |
| No contact/remote contact only | 2518 (77.8) | 2062 (77.6) | 456 (78.9) | Ref | - | Ref | - |
| Face to face but no physical contact | 149 (4.6) | 132 (5.0) | 17 (2.9) | 0.58 (0.35 – 0.97) | 0.04 | 0.60 (0.36 – 1.01) | 0.06 |
| Physical contact | 377 (11.7) | 312 (11.7) | 65 (11.3) | 0.94 (0.71 – 1.25) | 0.68 | 1.03 (0.77 – 1.38) | 0.84 |
| missing | 191 (5.9) | 151 (5.7) | 40 (6.9) |  |  |  |  |
| **Current or previous SARS-CoV-2 vaccine hesitancy** |  |  |  |  |  |  |  |
| Not hesitant | 2239 (69.2) | 1780 (67.0) | 459 (79.4) | Ref | - | Ref | - |
| Hesitant | 862 (26.7) | 725 (27.3) | 92 (15.9) | 0.49 (0.39 – 0.63) | <0.001 | 0.56 (0.43 – 0.71) | <0.001 |
| Missing | 134 (4.1) | 152 (5.7) | 27 (4.7) |  |  |  |  |
| **Number of influenza vaccines in previous 2 seasons** |  |  |  |  |  |  |  |
| 0 | 456 (14.1) | 397 (14.9) | 59 (10.2) | Ref | - | Ref | - |
| 1 | 515 (15.9) | 428 (16.1) | 87 (15.1) | 1.37 (0.96 – 1.96) | 0.09 | 1.35 (0.94 – 1.93) | 0.11 |
| 2 | 2123 (65.6) | 1717 (64.6) | 406 (70.2) | 1.59 (1.19 – 2.14) | 0.002 | 1.48 (1.10 – 1.99) | 0.01 |
| Missing | 141 (4.4) | 115 (4.3) | 26 (4.5) |  |  |  |  |
| **VAX score, med (IQR)** | 16 (14 – 17) | 16 (14 – 17) | 16 (15 – 18) | 1.11 (1.06 – 1.15) | <0.001 | 1.10 (1.06 – 1.14) | <0.001 |
| Missing | 0 (0.0) |  |  |  |  |  |  |
| **Number of comorbidities** |  |  |  |  |  |  |  |
| 0 | 2465 (76.2) | 2033 (76.5) | 432 (74.7) | Ref | - | Ref | - |
| ≥1 | 249 (7.7) | 201 (7.6) | 48 (8.3) | 1.12 (0.81 – 1.57) | 0.49 | 1.10 (0.79 – 1.55) | 0.56 |
| Missing | 521 (16.1) | 423 (15.9) | 98 (17.0) |  |  |  |  |
| **Pregnant\*** |  |  |  |  |  |  |  |
| Not pregnant | 2947 (91.1) | 2410 (90.7) | 537 (92.9) | Ref | - | Ref | - |
| Pregnant | 63 (2.0) | 58 (2.2) | 5 (0.9) | 0.39 (0.15 – 0.97) | 0.04 | 0.62 (0.24 – 1.56) | 0.31 |
| Missing | 225 (7.0) | 189 (7.1) | 36 (6.2) |  |  |  |  |
| **Previous COVID-19** |  |  |  |  |  |  |  |
| Never tested | 278 (8.6) | 229 (8.6) | 49 (8.5) | 1.00 (0.72 – 1.39) | 0.98 | 0.98 (0.70 – 1.36) | 0.88 |
| Tested negative | 2181 (67.4) | 1798 (67.7) | 383 (66.3) | Ref | - | Ref | - |
| Tested positive | 763 (23.6) | 620 (23.3) | 143 (24.8) | 1.08 (0.88 – 1.34) | 0.46 | 1.14 (0.92 – 1.40) | 0.25 |
| Missing | 13 (0.4) | 10 (0.4) | 3 (0.5) |  |  |  |  |
| **Lives with a person ≥65 years old** |  |  |  |  |  |  |  |
| No | 2808 (86.8) | 2314 (87.1) | 494 (85.5) | Ref | - | Ref | - |
| Yes | 320 (9.9) | 255 (9.6) | 65 (11.3) | 1.19 (0.89 – 1.59) | 0.23 | 0.88 (0.64 – 1.20) | 0.41 |
| Missing | 107 (3.3) | 88 (3.3) | 19 (3.3) |  |  |  |  |
| **Lives with other key workers** |  |  |  |  |  |  |  |
| No | 1678 (51.9) | 1378 (51.9) | 300 (51.9) | Ref | - | Ref | - |
| Yes | 1444 (44.6) | 1187 (44.7) | 257 (44.5) | 0.99 (0.83 – 1.20) | 0.95 | 1.03 (0.85 – 1.24) | 0.77 |
| Missing | 113 (3.5) | 92 (3.5) | 21 (3.6) |  |  |  |  |
| **Trusts employing organisation to deal with concern about unsafe clinical practice** |  |  |  |  |  |  |  |
| Does not trust organisation | 844 (26.1) | 673 (25.3) | 164 (28.4) | Ref | - | Ref | - |
| Trusts organisation | 2161 (66.8) | 1738 (65.4) | 350 (60.6) | 0.83 (0.67 – 1.02) | 0.07 | 0.78 (0.63 – 0.96) | 0.02 |
| Missing | 230 (7.1) | 246 (9.3) | 64 (11.1) |  |  |  |  |
| **COVID-19 conspiracies score, med (IQR)** | 8 (7 – 10) | 8 (7 – 10) | 8 (7 – 10) | 0.94 (0.89 – 0.98) | 0.004 | 0.95 (0.91 – 1.00) | 0.05 |
| Missing | 128 (4.0) | 107 (4.0) | 21 (3.6) |  |  |  |  |

\*multivariable logistic regression adjusted for age, sex and ethnicity
Data in the first two columns are n(%) with percentages computed column-wise apart from the total number of those favouring/not favouring mandatory vaccination which are computed row wise. Participants are coded as pregnant if they indicated they were pregnant at baseline or at follow up (i.e. this category includes participants who were pregnant at baseline and who may have delivered or any other reason leading to loss of pregnancy by the time of completing the follow up questionnaire). Due to including only complete cases in logistic regression models, multivariable models are based on fewer observations than univariable models. COVID-19 - coronavirus disease 2019; IMD – index of multiple deprivation; IQR – interquartile range; med – median; Ref – reference category for categorical variables in logistic regression analyses; SARS-CoV-2 – severe acute respiratory syndrome coronavirus-2

**Table 2:** Description of codes used to analyse free-text responses to the question “What should society do if people don’t get vaccinated against COVID-19?”

| Codes | Description of responses within code |
| --- | --- |
| Mandatory vaccination | Agreement with mandatory vaccination, either directly (e.g. by introducing a law) or via restrictions for unvaccinated individuals on access to essential services (work, education, healthcare), or by other severe restrictions such as isolating unvaccinated people completely and/or making them stay at home until they get vaccinated. |
| Mandatory vaccination for healthcare workers and social care staff | Agreement with mandatory vaccination for health and social care workers and those working with clinically vulnerable populations. |
| Specific restrictions for unvaccinated people | Vaccine passports and restricting access to leisure and travel but not to essential services (e.g. healthcare, work, education) and/or increased restrictions (e.g. wearing PPE, social distancing) for unvaccinated only. |
| Do nothing | Statements about freedom of choice being vital in a democracy, about it being immoral or undemocratic or against human rights to require people to be vaccinated. Statements that nothing can be done. |
| Educate, increase access or incentivise | Educate to improve understanding of the benefits and risks of vaccination. Encourage vaccine uptake (e.g. demonstrating empathy and listening). Providing positive incentives for vaccination (not restrictions on unvaccinated). Focus on improving uptake within marginalised groups by increasing confidence and knowledge (e.g. via relevant role models) and by tackling wider underlying societal causes of hesitancy (e.g. poverty and discrimination). Greater transparency about vaccine manufacture and roll-out. Improve access to vaccines or improving vaccines to make them more acceptable to some groups (e.g. vegans). |
| Maintain COVID-19 restrictions | Maintaining restrictions within society generally, not for unvaccinated people only (e.g. widespread use of PPE, social distancing, lockdowns). Consequences of higher levels of disease e.g. increased mortality within the population. |
| Don't know what to do | Statements about being unable to decide what to do e.g. “I really don't know”. Gives two opposing arguments (e.g. unable to do anything because of civil liberties, and mandatory vaccination for healthcare workers) without stating a preference for either. |
| Missing / not coded | Left blank. Unable to code because the response does not attempt to answer the question. |

#### Predictor variables

We chose the following predictor variables based on our previous analysis of the predictors of SARS-CoV-2 vaccine hesitancy^6^ together with a priori hypotheses about factors which might have an association with the outcome. These were as follows (see study protocol^27^ for details of each variable): demographics (ethnicity using Office for National Statistics categories^32^, age, sex, index of multiple deprivation (IMD) quintile of home postcode), migration status, job role, SARS-CoV-2 vaccine hesitancy as defined in our previous analysis^6^, influenza (flu) vaccine uptake in the previous two seasons, attitudes towards vaccination generally (VAX scale),^33^ presence of comorbidities, previous COVID-19, pregnancy, living with someone over the age of 65, living with other key workers, trust in employer organisation (to deal with a concern about unsafe clinical practice), number of confirmed/suspected COVID-19 patients seen per week, and belief in COVID-19 conspiracies.^34^

#### Statistical analysis

To assess response bias, we compared the demographic and occupational characteristics of those that responded to the free-text question with those that did not using chi-squared tests for categorical variable and Wilcoxon rank-sum tests for continuous variables.

We calculated the frequency and proportion of participants assigned to each code. We explored univariable relationships between predictor variables and the multinomial outcome variable using chi-squared tests for categorical variables and Kruskal-Wallis tests for continuous variables.

We used logistic regression to calculate the univariate association between the binary outcome variable and each predictor variable. We then calculated a base model by regressing age, sex and ethnicity on the binary outcome variable. We used logistic regression to examine the association of each of the other predictor variables in turn with the binary outcome, adjusted for age, sex and ethnicity.

We excluded participants with missing outcome data.

We used Stata 17 for all statistical analyses.^35^ P values <0.05 were considered statistically significant.

### Ethical approval

Both studies were approved by the Health Research Authority (Brighton and Sussex Research Ethics Committee; ethics reference: 20/HRA/4718). All participants gave written informed consent.

### Involvement and engagement

We worked closely with a Professional Expert Panel of HCWs from a range of ethnic backgrounds, occupations, and genders, as well as with national and local organisations (see study protocol).^27^

## Results

### Description of the analysed cohort

Of the 5633 respondents to both first and second questionnaires, 3235 (57.8%) gave a response to the free-text question that could be coded. Of these, 2405 (74.3%) were female and 897 (27.7%) were from ethnic minority groups. The majority of participants worked in allied health professional (40.3%), medical (23.9%) or nursing/midwifery (21.6%) roles (Table 1).

### Assessment of response bias

Supplementary Table 2 shows a comparison of the demographic and occupational characteristics of those that did and did not respond to the free text question. Non-responders were significantly younger than responders (median [IQR] age in years 43 [34 – 57] vs 47[36 – 56], p<0.001). There were no significant differences by sex, ethnicity or job role.

### Inter-rater reliability of coding

Of the 362 double-coded responses, the researchers agreed on 334 and disagreed on 28, giving a reliability score of 92.2%. Following discussion and resolution of all discrepancies, one code with very few responses (“Change vaccines”) was subsumed in the code “Educate, increase access, incentivise”.

### Qualitative description and frequency of codes

The analysis resulted in seven codes: Mandatory vaccination for the general population; Mandatory vaccination for HCWs; Do nothing; Specific restrictions for unvaccinated individuals; Maintain Covid-19 restrictions; Educate, increase access, incentivise; Don’t know. Codes are described briefly in Table 1. More detailed descriptions of each code with exemplar quotes and frequencies are given below.

#### Mandatory vaccination for the general population

Overall, 377 (12%) participants advocated mandatory vaccination for the general population and/or imposing serious sanctions on those who were eligible to be vaccinated but choose not to. This included fines and being shamed or isolated from society:

> *“Make it mandatory with meaningful penalties*.*”*
>
> *“Identify & Isolate them”*
>
> *“Shun them, and set them apart”*

A number felt that vaccination was necessary to relieve some of the pressure faced by HCWs and the NHS due to COVID-19, and suggested individuals who chose not to get vaccinated should be refused healthcare on the NHS and made to pay:

> *“People need to realise that they put health care workers at risk would sign a disclaimer for reduced treatment”*
>
> *“Make it mandatory or fine them, remove health services from them”*
>
> *“If people have capacity and refuse then they should relinquish their right to NHS treatment if they catch COVID 19. They should be made to pay for their treatment if they require hospital admission”*

Several also justified mandatory vaccination via the benefits to others and society from reducing the risk of transmission:

> *“Make them mandatory as part of our “social” and “national” responsibility to ourselves and others. Unless people cannot be vaccinated [sic] due to health reasons”*
>
> *“I hope everyone should consider getting vaccine to reduce the risk to themselves and others, or government should make it compulsory”*

#### Mandatory vaccination for HCW / social care staff

Overall, 201 (6%) participants specifically advocated mandatory vaccination for HCW and others working with vulnerable populations. Frequently this was described as an exceptional case that was necessary to protect the most vulnerable in society:

> *“Tough one. I think in certain roles this should be mandatory-healthcare workers, care home staff etc”*
>
> *“A really tricky one! I would support mandatory vaccination for people working in direct patient care roles (NHS/care sector) but not elsewhere”*

Several mentioned that vaccination against Hepatitis B was already required for HCWs:

> *“Depends on the reason for not getting vaccinated and what the individual is hoping to do. In healthcare roles, vaccination should be mandatory (c*.*f Hep B). In general society there should not be a compulsion”*

#### Specific restrictions for unvaccinated people

Overall, 17% (n=547) participants advocated for specific restrictions for unvaccinated individuals. Responses described restrictions on travel, social and leisure activities for those who are unvaccinated, including use of vaccine passports and additional testing or use of protective personal equipment:

> *“I think there should be a requirement for proof of vaccination or a negative covid test result for access to indoor event such as theatres, concerts etc and large scale outdoor events so that everyone can feel safe and confident. Those that refuse vaccination have the freedom and right to do so, but everyone has the right to feel safe”*
>
> *“Restrict access to social gatherings, travel etc if people choose not to get vaccinated”*

#### Do nothing

Overall 18% (n=580) had a response coded as “Do nothing”; a similar proportion as advocated mandatory vaccination. Common reasons in this category referred to the belief that the decision to be vaccinated was a personal choice that it was important to respect:

> *“Accept that this is a free country and people are capable of making their own decisions” “No idea. Very hard to do as it is a choice and against human rights to enforce it”*
>
> *“People should have a choice. They should not feel pressured into having something we still do not know a lot about*.*”*
>
> *“It’s a personal choice. We cannot force people to inject themselves with a substance. I think this could be considered as assault if we were made to*.*”*

In contrast to the participants who felt vaccination should be mandatory to protect others, several participants in this category stated their opinion that unvaccinated people were only putting themselves at risk:

> *“Nothing - carry on. if people want the vaccine then that it ok but it is also ok not to have this. the vaccine reduces the symptoms if you get covid, it doesn’t stop you getting it or transmitting it*.*”*
>
> *“Nothing much. If someone wishes to risk their life by not getting vaccinated, society cannot prevent this. Same applies to dangerous activities like mountaineering horse-riding etc*.*”*

A few described how other vaccinations were not mandatory, so there was little justification for making Covid-19 vaccinations compulsory:

> *“People decline the flu vaccine and face no repercussions so I don’t see how any sanctions can be placed on people not having the covid vaccine”*.

Some feared that making vaccination mandatory could have negative consequences:

> *“Best to careful - there are risks of creating a two tier society or driving the non-vaccinated underground”*
>
> *“Just carry on and try not to use it as an excuse to argue and build bigger barriers”*

A few mentioned that some people are clinically unable to be vaccinated:

> *“Respect people’s wishes as it may be no fault of their own eg health or capacity reasons”*

Several mentioned that herd immunity, either from vaccination or infection would be sufficient for tackling COVID-19:

> *“Allow choice, hope herd immunity overcomes”*
>
> *“Return to normal as large numbers and vulnerable have been vaccinated*.*”*

One participant stated a preference for “natural” immunity over vaccination:

> *“Cheer. Most people have a robust immune system. We will have been exposed to the usual coronaviruses/rhinoviruses during our lifetime and our t-cells and lymphocytes will recognise these again. Those with co morbidities should shield if they wish*.*”*

#### Educate, increase access or incentivise

The most frequent code, chosen by almost a third of participants overall (1047/3229, 32%) was to educate, increase access to vaccines or incentivise vaccination. Many responses described the need to provide better information and support that would enable people to make decisions based on evidence and an assessment of the risks and benefits:

> *“Continue to encourage; ensure people aware of the evidence for vaccine and consequences of not doing it”*

Several responses specifically mentioned the role of religious and ethnic minority community leaders in encouraging vaccination:

> *“Work with them and find out why they do not want it. Enlist community champions, eg religious leaders or other community leaders”*
>
> *“Further support through campaigns, education and use of local champions or champions from same ethnicity*.*”*

There were some calls for those spreading or allowing the spread of false information to be sanctioned:

> *“Media and social media should be held accountable for spreading false information”*

#### Maintain COVID-19 restrictions

Overall, 10% (336) felt that society’s response to suboptimal vaccination coverage would be to maintain COVID-19 restrictions, including for those shielding or especially vulnerable.

> *“Remain on specific lock downs or continue with strict social distancing measure[s]”*

For some participants this may have been seen as a solution, but for others it reflected a feeling of resignation:

> *“More lockdowns will be inevitable” “Prepare for another wave of infection!”*

#### Don’t know

A minority (147, 5%) acknowledged this is a difficult a question to answer or didn’t give a definitive answer:

> *“It’s a difficult one. Everyone has a right to decline, but this risks the greater good for everyone. These people will get herd immunity. I don’t have a real answer for this dilemma*.*”*

### Statistical associations of predictor variables with outcome measures

Overall 18% (n=578) favoured mandatory vaccination (either in general or for HCWs specifically), whereas 82% favoured other options. Table 1 shows the cohort stratified by the binary outcome variable and by the predictor variables with p values from univariable and multivariable tests of association. Supplementary Table 1 shows the cohort stratified by the seven-level categorical outcome variable and by the predictor variables with p values from univariable tests of association.

Univariable analyses indicated that favouring mandatory vaccination was associated with being older (OR 1.29; 95%CI 1.19 – 1.39, per decade increase), having been vaccinated against influenza in both the previous two seasons (OR 1.59; 95%CI 1.19 – 1.24, vs none), and with more pro-vaccine attitudes (OR 1.11; 95%CI 1.06 – 1.15); whereas not favouring mandatory vaccination was associated with female sex (OR 0.78; 95%CI 0.64 – 0.95, vs male), Black (OR 0.46; 95%CI 0.25 – 0.85, vs white) or Asian (OR 0.75; 95%CI 0.59 – 0.97, vs white) ethnicity, Allied Health Professional occupation (OR 0.70; 95%CI 0.54 – 0.90, vs Medical), hesitancy about SARS-CoV-2 vaccination (OR 0.49; 95%CI 0.39 – 0.64, vs not hesitant), pregnancy (current or recent, 0.39; 95%CI 0.15 – 0.97, vs not pregnant) and greater belief in COVID-19 conspiracies (OR 0.94; 95%CI 0.91 – 1.00).

Multivariable analysis from the base model (Figure 1a) showed that older HCWs were more likely to favour mandatory vaccination (OR 1.26; 95%CI 1.17 – 1.37, per decade increase) whereas female HCWs (OR= 0.80, 95%CI 0.65 – 0.99, vs male) and HCWs of Black ethnicity (OR= 0.48, 95%CI 0.26 – 0.87, vs White) were less likely to. After adjusting for age, sex and ethnicity (Figure 1b), HCWs vaccinated against influenza in the previous two seasons (OR 1.48; 95%CI 1.10 – 1.99, vs no influenza vaccine) and with more positive attitudes towards vaccination generally (OR 1.10; 95%CI 1.06 – 1.14) were more likely to favour mandatory vaccination, whereas those hesitant about COVID-19 vaccination (OR= 0.56; 95% CI 0.43 – 0.71, vs not hesitant), who were in an Allied Health Profession (OR 0.67; 95%CI 0.51 – 0.88, vs Medical), or who trusted their organisation to act on a concern they raised about unsafe clinical practice (OR 0.78; 95%CI 0.63 – 0.96) were less likely to favour mandatory vaccination.

**Figure 1.**
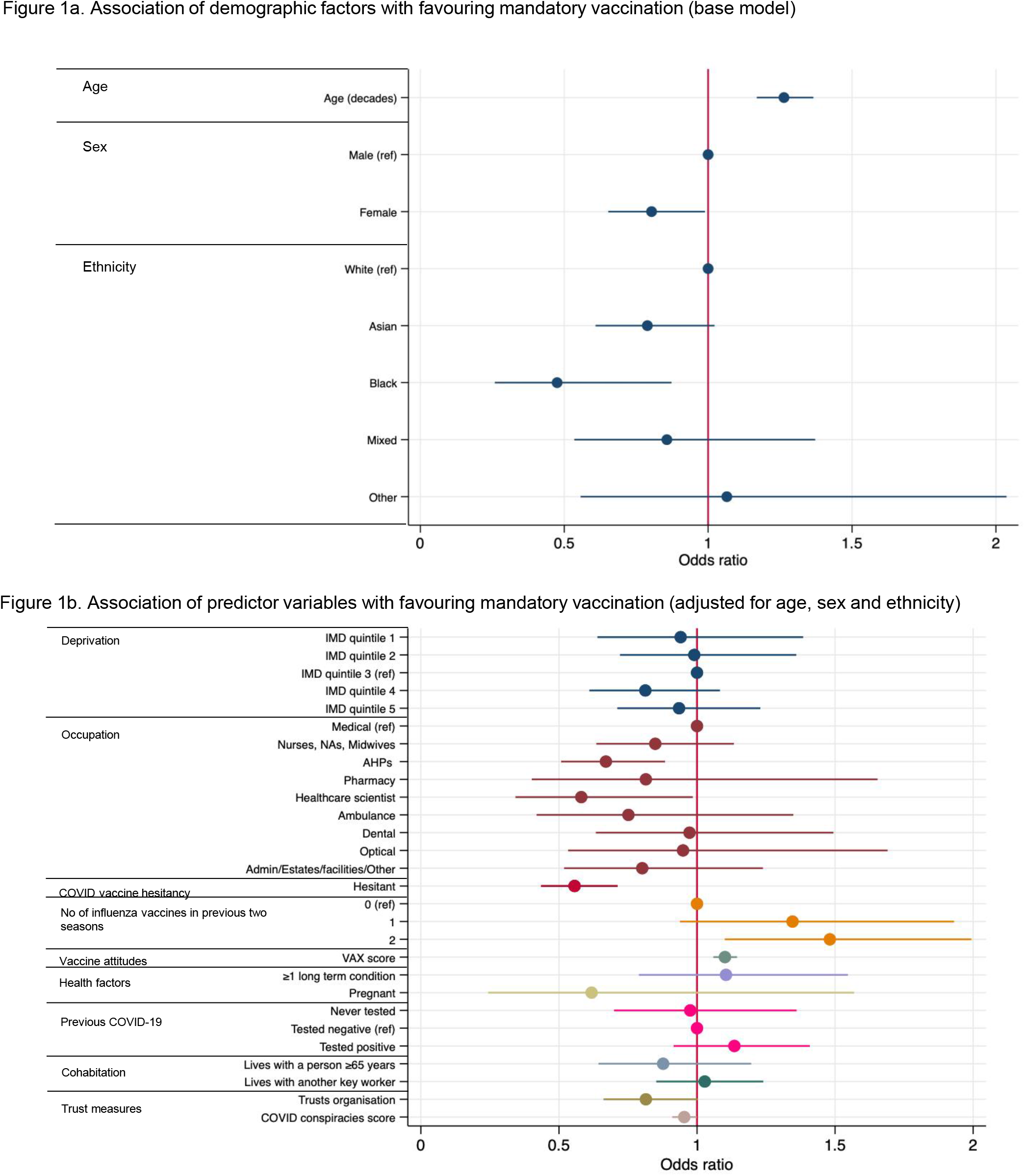
Fig 1a shows the results of multivariable logistic regression. Associations of age, sex and ethnicity with an outcome of favouring mandatory vaccination are expressed as adjusted odds ratios and 95% confidence intervals. Regression analyses contained complete cases only, therefore, this analysis includes 3215 HCW. Fig 1b shows the results of multivariable logistic regression. Associations of predictor variables (adjusted for age, sex and ethnicity) with an outcome of favouring mandatory vaccination are expressed as adjusted odds ratios and 95% confidence intervals. Regression analyses contained complete cases only, therefore, estimates for each predictor variable are based on different numbers of observations. Sample size for each predictor can be calculated by subtracting the number of observations with missing data (table 2) for the predictor from the number of observations in the base model (n=3215). AHP – allied health professional; COVID-19 – coronavirus disease 2019; IMD – index of multiple deprivation

## Discussion

### Summary of results

Only one in six HCWs in this large, diverse, UK-wide sample favoured mandatory vaccination as a strategy for dealing with suboptimal vaccination coverage; a third favoured education and support as a strategy. HCWs who were hesitant about vaccination generally and about COVID-19 vaccination specifically, as well as female HCWs, HCWs from some ethnic minority groups, younger HCWs, and Allied Health Professional occupational groups were less likely to advocate mandatory vaccination.

### Strengths and limitations

This is to our knowledge, the only large-scale national study of HCW perceptions of mandatory vaccination. The size and the diversity of the sample, together with detailed information collected on participants from two waves of the UK-REACH longitudinal questionnaire, meant it was possible to examine differences in views by participant ethnicity, occupational group, age, health, and vaccination status.

Responses were collected in early Summer 2021, before mandatory vaccination for health and social care staff was announced in England on 9^th^ November 2021,^12^ and before the classification by the UK Health Security Agency of the Omicron variant of SARS-CoV-2 as a variant of concern on 27^th^ November 2021^36^ with greater transmissibility. This may have altered attitudes towards mandatory vaccination and vaccine passports. Data collection did however coincide with the classification by the UK Government of the Delta variant as a variant of concern on 7^th^ May 2021, with the roll-out of vaccines to all eligible adults in the UK by July 2021, and the lifting of restrictions in June 2021 after a third national lockdown.^37^ Indeed, since the start of the vaccine roll-out increasing data on the vaccines has continued to emerge may have influenced and continue to influence participants’ views about mandatory vaccination in a variety of ways. This includes data on rare-side effects of the vaccines,^38,39^ the effectiveness of Covid-19 vaccination in pregnancy,^40^ the high efficacy of the vaccines against severe disease and hospitalisation^41^ and the impact of vaccination on reducing household transmission of Covid-19.^42^

As with any questionnaire study, there is the potential for selection bias, although the baseline UK-REACH study population is broadly representative of the NHS workforce, albeit with a lower proportion of ancillary staff.^6^ The outcome variable data were gathered via a free-text question and we were therefore unable to explore attitudes and opinions in as much depth as would have been possible in a qualitative interview study. Our use of a free-text question rather than a multiple-choice question meant we could not capture what each participant thought about each code, including mandatory vaccination, however we were able to capture information that is potentially representative of participants’ priorities since they generated the responses themselves. The participant-generated responses to the free-text question also allowed us to capture new information about HCWs’ views not necessarily covered in the literature; however we have included a quantitative question about mandatory vaccination in a subsequent questionnaire. The trustworthiness of the results is also supported by the high inter-rater reliability of the double-coding of a large portion of responses. Our method of only allowing one code per response, and of choosing the most socially-restrictive code when there were multiple options, meant we did not examine the co-occurrence of codes within participants. This could also have resulted in an under-estimation of the endorsement of less restrictive options, such as Do nothing. The fact that respondents to the free-text question were slightly older than the cohort generally may have resulted in the over-estimation of the endorsement of more restrictive options such as mandatory vaccination for HCWs, which were more common among older participants.

### Comparisons with other research

Other studies have assessed HCW views on mandatory vaccination for non-COVID diseases, finding mixed levels of support. For example, a systematic review and meta-analysis of studies of HCW views on mandatory influenza vaccination found 61% agreeing with mandatory vaccination, but agreement varied between studies considerably, ranging from 15% to 90%. Rates of agreement were higher among vaccinated HCWs and among doctors (physicians and general practitioners compared to nurses).^24^ A study of medical students at one Austrian university published in 2020 found 83% agreed with mandatory hepatitis B vaccination, whereas only 40% agreed with mandatory influenza vaccination.^25^ A survey of English healthcare staff responsible for influenza vaccination campaigns conducted in 2017 found that 68% believed a mandatory campaign would increase uptake, but only 17% of those surveyed believed other HCWs would support mandatory vaccination, and qualitative interview and focus group results demonstrated “fairly consistent opposition” to mandatory vaccination.^43^ In relation to COVID-19 vaccination, a mixed-methods study of health and social care workers published as a pre-print found that participants who agreed that they felt under pressure from their employer to get a vaccine were more likely to have declined vaccination, although the direction of causality is unclear.^44^

### Policy and practice implications, and future research

Only 12% of HCWs in our study proposed mandatory COVID-19 vaccination for the general public as a solution to sub-optimal vaccination rates, and half that number (6%) specifically proposed mandatory vaccination for HCWs. This suggests that the policy to introduce mandatory vaccination for frontline HCWs in England could be unpopular with the majority of HCWs, especially those who are already hesitant about vaccination against COVID-19 or against other diseases, as well as among female HCWs who make up the majority of NHS staff, and among Black and Asian ethnic groups as well as among Allied Health Professionals. If the introduction of mandatory vaccination does reduce trust in healthcare providers and employers among those groups, this could exacerbate existing health inequalities, and could be perceived as counteracting the ethical principle that individuals should be able to give true informed consent to a medical intervention.^45^

It is vital that strong efforts are made to build confidence in the safety and efficacy of vaccines among those who are hesitant.^46^ This includes vaccines against COVID-19, as well as vaccines against influenza and other vaccine-preventable diseases that compromise NHS staff health and patient safety. Building vaccine confidence among HCWs will not just increase uptake, but also enable HCWs to advocate for vaccination among patients^47^ and members of their own communities. This may support uptake among groups experiencing social and health inequalities who may be at greater risk of adverse effects from COVID-19 as well as other diseases.^48^

The findings of this study cast some doubt on the potential success of a mandatory vaccination policy for HCWs. The UK Government’s own impact assessment for its proposed mandatory vaccination policy estimates that only a minority of currently unvaccinated HCWs would be vaccinated under the policy, which would leave 88,000 HCWs (5% of the workforce) unvaccinated and non-exempt, and therefore needing to be removed from patient-facing roles.^12^ Meanwhile the King’s Fund has stated that NHS hospitals, mental health services and community providers reported a shortage of nearly 84,000 full-time equivalent staff in 2020, which might be exacerbated by enforcement of sanctions against unvaccinated HCWs^49^.

Future research should include evaluation of specific approaches for increasing vaccine uptake among HCWs, their colleagues, patients, families and communities. UK-REACH is currently analysing in-depth qualitative data on vaccine perceptions, and collecting longitudinal data on attitudes to vaccination policies and changes in vaccine hesitancy and uptake among diverse HCWs, which will help inform the best approaches to maximise uptake in this vital group.

## Conclusions

The majority of UK HCWs did not propose mandatory Covid-19 vaccination as a solution to sub-optimal vaccine coverage. Mandatory vaccination was less favoured by those already hesitant about vaccination against Covid-19 and influenza, and among HCWs who are women, from some ethnic minority groups, younger, and in Allied Health Profession occupations. While compulsory vaccination is likely to increase coverage, a significant number of HCWs could remain unvaccinated and further research is required to understand the impact of compulsory vaccination policies in England, particularly on levels of trust among some minoritised groups, staff well-being, and shortages.

## Data Availability

To access data or samples produced by the UK-REACH study, the working group representative must first submit a request to the Core Management Group by contacting the UK-REACH Project Manager in the first instance. For ancillary studies outside of the core deliverables, the Steering Committee will make final decisions once they have been approved by the Core Management Group. Decisions on granting the access to data/materials will be made within eight weeks. Third party requests from outside the Project will require explicit approval of the Steering Committee once approved by the Core Management Group. Note that should there be significant numbers of requests to access data and/or samples then a separate Data Access Committee will be convened to appraise requests in the first instance.

https://www.uk-reach.org/main/data-dictionary/

## Contributors

MP conceived of the idea for the study and led the application for funding with input from MDT, KK, ICM, KW, RF, LBN, SC, KRA, LJG, ALG and CJ. The survey was designed by KW, MP, ICM, CMel, CJ, ALG, LBN, RF and CAM. Online consent and survey tools were developed by LB. The underlying data were verified by CAM, KW, MG and MP. KW, MG and CAM, wrote the first draft of the manuscript with input from MP, PP, and all co-authors. All authors should confirm that they had full access to all the data in the study and accept responsibility to submit for publication. All authors approved the submitted manuscript.

## Acknowledgements

We would like to thank all the healthcare workers who took part in this study when the NHS was under immense pressure.

We wish to acknowledge the Professional Expert Panel group and the Steering and Advisory Group (see the cohort study protocol^27^ for details), and SERCO, as well as the following people for their support in setting up the study from the regulatory bodies: Kerrin Clapton and Andrew Ledgard (General Medical Council), Caroline Kenny (Nursing and Midwifery Council), David Teeman and Lisa Bainbridge (General Dental Council), My Phan and John Tse (General Pharmaceutical Council), Angharad Jones (General Optical Council), Katherine Timms and Charlotte Rogers (The Health and Care Professions Council) and Mark Neale (Pharmaceutical Society of Northern Ireland).

We would also like to acknowledge the following trusts and sites who recruited participants to the study: Nottinghamshire Healthcare NHS Foundation Trust, University Hospitals Leicester, Lancashire Teaching Hospitals NHS Foundation Trust, Northumbria Healthcare, Berkshire Healthcare, Derbyshire Healthcare NHS Foundation Trust, South Tees NHS Foundation Trust, Birmingham and Solihull NHS Foundation Trust, Affinity Care, Royal Brompton and Harefield, Sheffield Teaching Hospitals, St George’s Hospital, Yeovil District Hospital, Lewisham and Greenwich NHS Trust, Black Country Community Healthcare NHS Foundation Trust, Sussex Community NHS Foundation Trust, South Central Ambulance Service, University Hospitals Coventry and Warwickshire, University Hospitals Southampton NHS Foundation Trust, London Ambulance Trust, Royal Free, Birmingham Community Healthcare NHS Foundation Trust, Central London Community Healthcare, Chesterfield Royal Hospital, Bridgewater Community Healthcare, Northern Borders, County Durham and Darlington Foundation Trust, Walsall Healthcare NHS Trust.

## Transparency statement

The lead authors affirm that this manuscript is an honest, accurate, and transparent account of the study being reported; that no important aspects of the study have been omitted; and that any discrepancies from the study as planned (and, if relevant, registered) have been explained.

## Disclaimers

The views expressed in the publication are those of the author(s) and not necessarily those of the National Health Service (NHS), the NIHR or the Department of Health and Social Care. This research was funded in whole, or in part, by the Wellcome Trust [WT204801/Z/16/Z and WT 202849/Z/16/Z]. For the purpose of open access, the author has applied a CC BY public copyright licence to any Author Accepted Manuscript version arising from this submission.

---

**Figure 1 shows the multivariable logistic regression results.**

Figure 1a shows the base model. Associations of age, sex and ethnicity with an outcome of favouring mandatory vaccination are expressed as adjusted odds ratios and 95% confidence intervals. Regression analyses contained complete cases only, therefore, this analysis includes 3215 HCW.

Figure 1b shows the associations of predictor variables (adjusted for age, sex and ethnicity) with an outcome of favouring mandatory vaccination. Results are expressed as adjusted odds ratios and 95% confidence intervals. Regression analyses contained complete cases only, therefore, estimates for each predictor variable are based on different numbers of observations. Sample size for each predictor can be calculated by subtracting the number of observations with missing data (See Table 1) for the predictor from the number of observations in the base model (n=3215).

AHP – allied health professional; COVID-19 – coronavirus disease 2019; IMD – index of multiple deprivation

