## Supplementary Table 1 for "Healthcare workers’ views on mandatory SARS-CoV-2 vaccination in the United Kingdom: findings from the UK-REACH prospective longitudinal cohort study"

**Supplementary Table 1. Description of the cohort stratified by coded response to the free text question and selected predictors**

| Variable | Total | Code | | | | | | | P value† |
|---|---|---|---|---|---|---|---|---|---|
| | | Do nothing | Educate, increase access or incentivise | Maintain restrictions | Specific restrictions for unvaccinated | Mandatory vaccination for HCW / social care staff | Mandatory vaccination for general population or limited access to vital services | Don't know | |
| | **N=3235** | **580 (17.9)** | **1047 (32.4)** | **336 (10.4)** | **547 (16.9)** | **201 (6.2)** | **377 (11.7)** | **147 (4.5)** | |
| **Age**, med(IQR) | 46 (35 – 55) | 46 (36 – 56) | 48 (38 – 56) | 41 (32 – 52) | 46 (35 – 56) | 54 (46 – 61) | 49 (38 – 57.5) | 46 (35 – 56) | <0.001 |
| Missing | 13 (0.4) | 6 (1.0) | 2 (0.2) | 0 (0.0) | 2 (0.4) | 2 (1.0) | 1 (0.3) | 0 (0.0) | |
| **Sex** | | | | | | | | | |
| Male | 824 (25.5) | 132 (22.8) | 292 (27.9) | 65 (19.4) | 138 (25.2) | 43 (21.4) | 128 (34.0) | 26 (17.7) | <0.001 |
| Female | 2405 (74.3) | 445 (76.7) | 754 (72.0) | 271 (80.7) | 408 (74.6) | 158 (78.6) | 248 (65.8) | 121 (82.3) | |
| Missing | 6 (0.2) | 3 (0.5) | 1 (0.1) | 0 (0.0) | 1 (0.2) | 0 (0.0) | 1 (0.3) | 0 (0.0) | |
| **Ethnicity** | | | | | | | | | |
| White | 2336 (72.2) | 444 (76.6) | 688 (65.7) | 231 (68.8) | 411 (75.1) | 170 (84.6) | 275 (72.9) | 117 (79.6) | <0.001 |
| Asian | 571 (17.7) | 78 (13.5) | 236 (22.5) | 71 (21.1) | 84 (15.4) | 16 (8.0) | 70 (18.6) | 16 (10.9) | |
| Black | 122 (3.8) | 28 (4.8) | 52 (5.0) | 10 (3.0) | 16 (2.9) | 3 (1.5) | 9 (2.4) | 4 (2.7) | |
| Mixed | 144 (4.5) | 26 (4.5) | 47 (4.5) | 15 (4.5) | 27 (4.9) | 7 (3.5) | 16 (4.2) | 6 (4.1) | |
| Other | 60 (1.9) | 3 (0.5) | 24 (2.3) | 9 (2.7) | 9 (1.7) | 5 (2.5) | 7 (1.9) | 3 (2.0) | |
| Missing | 2 (0.1) | 1 (0.2) | 0 (0.0) | 0 (0.0) | 0 (0.0) | 0 (0.0) | 0 (0.0) | 1 (0.7) | |
| **Migration status** | | | | | | | | | |
| Born in the UK | 2356 (72.8) | 447 (77.1) | 709 (67.7) | 233 (69.4) | 424 (77.5) | 164 (81.6) | 267 (70.8) | 112 (76.2) | <0.001 |
| Born outside the UK | 791 (24.5) | 120 (20.7) | 306 (29.2) | 91 (27.1) | 114 (20.8) | 33 (16.4) | 101 (26.8) | 26 (17.7) | |
| Missing | 88 (2.7) | 13 (2.2) | 32 (3.1) | 12 (3.6) | 9 (1.7) | 4 (2.0) | 9 (2.4) | 9 (6.1) | |
| **IMD quintile** | | | | | | | | | |
| 1 (most deprived) | 275 (8.5) | 55 (9.5) | 88 (8.4) | 42 (12.5) | 33 (6.0) | 9 (4.5) | 36 (9.6) | 12 (8.2) | 0.002 |
| 2 | 472 (14.6) | 102 (17.6) | 147 (14.0) | 53 (15.8) | 60 (11.0) | 29 (14.4) | 55 (14.6) | 26 (17.7) | |
| 3 | 588 (18.2) | 99 (17.1) | 193 (18.4) | 59 (17.6) | 96 (17.6) | 36 (17.9) | 78 (20.7) | 27 (18.4) | |
| 4 | 727 (22.5) | 117 (20.2) | 243 (23.2) | 78 (23.2) | 136 (24.9) | 36 (17.9) | 84 (22.3) | 33 (22.5) | |
| 5 (least deprived) | 808 (25.0) | 124 (21.4) | 283 (27.0) | 67 (19.9) | 149 (27.2) | 68 (33.8) | 86 (22.8) | 31 (21.1) | |

| | | | | | | | | | |
|---|---|---|---|---|---|---|---|---|---|
| Missing | 365 (11.3) | 83 (14.3) | 93 (8.9) | 37 (11.0) | 73 (13.4) | 23 (11.4) | 38 (10.1) | 18 (12.2) | |
| **Job role** | | | | | | | | | |
| Medical | 773 (23.9) | 95 (16.4) | 301 (28.8) | 63 (18.8) | 133 (24.3) | 55 (27.4) | 99 (26.3) | 27 (18.4) | <0.001 |
| Nursing (inc Midwives + HCAs) | 698 (21.6) | 139 (24.0) | 196 (18.7) | 75 (22.3) | 115 (21.0) | 52 (25.9) | 82 (21.8) | 39 (26.5) | |
| AHPs* | 1303 (40.3) | 247 (42.6) | 416 (39.7) | 155 (46.1) | 218 (39.9) | 65 (32.3) | 135 (35.8) | 67 (45.6) | |
| Dental | 173 (5.4) | 38 (6.6) | 45 (4.3) | 21 (6.3) | 27 (4.9) | 14 (7.0) | 22 (5.8) | 6 (4.1) | |
| Administrative/estates/other | 195 (6.0) | 42 (7.2) | 58 (5.5) | 16 (4.8) | 39 (7.1) | 10 (5.0) | 24 (6.4) | 6 (4.1) | |
| Missing | 93 (2.9) | 19 (3.3) | 31 (3.0) | 6 (1.8) | 15 (2.7) | 5 (2.4) | 15 (4.0) | 2 (1.4) | |
| **Exposure to patients with COVID-19 (at time of second questionnaire)** | | | | | | | | | |
| No contact/remote contact only | 2518 (77.8) | 458 (79.0) | 809 (77.3) | 249 (74.1) | 431 (78.8) | 165 (82.1) | 291 (77.2) | 115 (78.2) | 0.26 |
| Face to face but no physical contact | 149 (4.6) | 29 (5.0) | 59 (5.6) | 19 (5.7) | 23 (4.2) | 6 (3.0) | 11 (2.9) | 2 (1.4) | |
| Physical contact | 377 (11.7) | 68 (11.7) | 116 (11.1) | 43 (12.8) | 65 (11.9) | 16 (8.0) | 49 (13.0) | 20 (13.6) | |
| missing | 191 (5.9) | 25 (4.3) | 63 (6.0) | 25 (7.4) | 28 (5.1) | 14 (7.0) | 26 (6.9) | 10 (6.8) | |
| **SARS-CoV-2 vaccine hesitancy** | | | | | | | | | |
| Not hesitant | 2239 (69.2) | 333 (57.4) | 750 (71.6) | 206 (61.3) | 412 (75.3) | 174 (86.6) | 285 (75.6) | 79 (53.7) | <0.001 |
| Hesitant | 862 (26.7) | 236 (40.7) | 244 (23.3) | 112 (33.3) | 116 (21.2) | 20 (10.0) | 78 (20.7) | 56 (38.1) | |
| Missing | 134 (4.1) | 11 (1.9) | 53 (5.1) | 18 (5.4) | 19 (3.5) | 7 (3.5) | 14 (3.7) | 12 (8.2) | |
| **Number of influenza vaccines in previous 2 seasons** | | | | | | | | | |
| 0 | 456 (14.1) | 119 (20.5) | 142 (13.6) | 48 (14.3) | 64 (11.7) | 16 (8.0) | 43 (11.4) | 24 (16.3) | <0.001 |
| 1 | 515 (15.9) | 100 (17.2) | 174 (16.6) | 54 (16.1) | 84 (15.4) | 30 (14.9) | 57 (15.1) | 16 (10.9) | |
| 2 | 2123 (65.6) | 342 (59.0) | 682 (65.1) | 218 (64.9) | 382 (69.8) | 147 (73.1) | 259 (68.7) | 93 (63.3) | |
| Missing | 141 (4.4) | 19 (3.3) | 49 (4.7) | 16 (4.8) | 17 (3.1) | 8 (4.0) | 18 (4.8) | 18 (4.8) | |
| **VAX score,** med (IQR) | 16 (14 – 17) | 15 (13 – 17) | 16 (14 – 17) | 15 (13 – 17) | 16 (15 – 17) | 16 (15 – 18) | 16 (14 – 18) | 16 (13 – 17) | <0.001 |
| Missing | 0 (0.0) | 0 (0.0) | 0 (0.0) | 0 (0.0) | 0 (0.0) | 0 (0.0) | 0 (0.0) | 0 (0.0) | |
| **Number of comorbidities** | | | | | | | | | |
| 0 | 2465 (76.2) | 451 (77.8) | 810 (77.4) | 258 (76.8) | 412 (75.3) | 147 (73.1) | 285 (75.6) | 102 (69.4) | 0.9 |
| ≥1 | 249 (7.7) | 43 (7.4) | 78 (7.5) | 27 (8.0) | 41 (7.5) | 20 (10.0) | 28 (7.4) | 12 (8.2) | |
| Missing | 521 (16.1) | 86 (14.8) | 159 (15.2) | 51 (15.2) | 94 (17.2) | 34 (16.9) | 64 (17.0) | 33 (22.5) | |
| **Pregnant**[‡] | | | | | | | | | |
| Not pregnant | 2947 (91.1) | 539 (92.9) | 938 (89.6) | 301 (89.6) | 505 (92.3) | 187 (93.0) | 350 (92.8) | 127 (86.4) | 0.16 |

| | | | | | | | | | |
|---|---|---|---|---|---|---|---|---|---|
| Pregnant | 63 (2.0) | 12 (2.1) | 20 (1.9) | 12 (3.6) | 12 (2.2) | 3 (1.5) | 2 (0.5) | 2 (1.4) | |
| Missing | 225 (7.0) | 29 (5.0) | 89 (8.5) | 23 (6.9) | 30 (5.5) | 11 (5.5) | 25 (6.6) | 18 (12.2) | |
| **Previous COVID-19** | | | | | | | | | |
| Never tested | 278 (8.6) | 55 (9.5) | 99 (9.5) | 31 (9.2) | 37 (6.8) | 21 (10.5) | 28 (7.4) | 7 (4.8) | 0.22 |
| Tested negative | 2181 (67.4) | 380 (65.5) | 708 (67.6) | 238 (70.8) | 376 (68.7) | 127 (63.2) | 256 (67.9) | 96 (65.3) | |
| Tested positive | 763 (23.6) | 141 (24.3) | 237 (22.6) | 65 (19.4) | 133 (24.3) | 51 (25.4) | 92 (24.4) | 44 (29.9) | |
| Missing | 13 (0.4) | 4 (0.7) | 3 (0.3) | 2 (0.6) | 1 (0.2) | 2 (1.0) | 1 (0.3) | 0 (0.0) | |
| **Lives with a person ≥65 years old** | | | | | | | | | |
| No | 2808 (86.8) | 510 (87.9) | 902 (86.2) | 294 (87.5) | 483 (88.3) | 166 (82.6) | 328 (87.0) | 125 (85.0) | 0.25 |
| Yes | 320 (9.9) | 56 (9.7) | 108 (10.3) | 29 (8.6) | 52 (9.5) | 30 (14.9) | 35 (9.3) | 10 (6.8) | |
| Missing | 107 (3.3) | 14 (2.4) | 37 (3.5) | 13 (3.9) | 12 (2.2) | 5 (2.5) | 14 (3.7) | 12 (8.2) | |
| **Lives with other key workers** | | | | | | | | | |
| No | 1678 (51.9) | 287 (49.5) | 560 (53.5) | 168 (50.0) | 283 (51.7) | 118 (58.7) | 182 (48.3) | 80 (54.4) | 0.05 |
| Yes | 1444 (44.6) | 279 (48.1) | 450 (43.0) | 155 (46.1) | 250 (45.7) | 74 (36.8) | 183 (48.5) | 53 (36.1) | |
| Missing | 113 (3.5) | 14 (2.4) | 37 (3.5) | 13 (3.9) | 14 (2.6) | 9 (4.5) | 12 (3.2) | 14 (9.5) | |
| **Trusts employing organisation to deal with concern about unsafe clinical practice** | | | | | | | | | |
| Does not trust organisation | 844 (26.1) | 165 (28.5) | 248 (23.7) | 88 (26.2) | 149 (27.2) | 45 (22.4) | 116 (30.8) | 33 (22.5) | 0.07 |
| Trusts organisation | 2161 (66.8) | 384 (66.2) | 724 (69.2) | 223 (66.4) | 361 (66.0) | 138 (68.7) | 230 (61.0) | 101 (68.7) | |
| Missing | 230 (7.1) | 31 (5.3) | 75 (7.2) | 25 (7.4) | 37 (6.8) | 18 (9.0) | 31 (8.2) | 13 (8.8) | |
| **COVID-19 conspiracies score,** med (IQR) | 8 (7 – 10) | 9 (8 – 11) | 8 (7 – 10) | 9 (8 – 10) | 8 (7 – 10) | 8 (7 – 9) | 9 (7 – 10) | 9 (8 – 10) | <0.001 |
| Missing | 128 (4.0) | 17 (2.9) | 44 (4.2) | 16 (4.8) | 16 (2.9) | 6 (3.0) | 15 (4.0) | 14 (9.5) | |

Also includes pharmacists, healthcare scientists, ambulance workers and those in optical roles. † groups compared using chi-squared tests for categorical variables and Kruskal-Wallis tests for continuous variables. Values expressed as n(%) unless stated otherwise. ‡ Participants are coded as pregnant if they indicated they were pregnant at baseline or at follow up (i.e. this category includes participants who were pregnant at baseline and who may have delivered by the time of completing the follow up questionnaire. IMD – index of multiple deprivation; IQR – interquartile range;
