## Supplementary Table 2 for "Healthcare workers’ views on mandatory SARS-CoV-2 vaccination in the United Kingdom: findings from the UK-REACH prospective longitudinal cohort study"

**Supplementary Table 2. Comparing demographic and occupational characteristics of free text responders and non-responders**

| | **Total**<br>**N=5633** | **Responders**<br>**N=3235** | **Non-responders**<br>**N=2398** | **P value*** |
|---|---|---|---|---|
| **Age, med(IQR)** | 46 (35 – 55) | 47 (36 – 56) | 43 (34 – 53) | <0.001 |
| **Sex** | | | | |
| Male | 1411 (25.1) | 824 (25.5) | 587 (24.5) | 0.38 |
| Female | 4215 (74.8) | 2405 (74.3) | 1810 (75.5) | |
| Missing | 7 (0.1) | 6 (0.2) | 1 (0.0) | |
| **Ethnicity** | | | | |
| White | 4,106 (72.9) | 2336 (72.2) | 1770 (73.8) | 0.32 |
| Asian | 984 (17.5) | 571 (17.7) | 413 (17.2) | |
| Black | 197 (3.5) | 122 (3.8) | 75 (3.1) | |
| Mixed | 236 (4.2) | 144 (4.5) | 92 (3.8) | |
| Other | 103 (1.8) | 60 (1.9) | 43 (1.8) | |
| Missing | 7 (0.1) | 2 (0.1) | 5 (0.2) | |
| **Job role** | | | | |
| Medical | 1365 (24.2) | 778 (24.1) | 587 (24.5) | 0.22 |
| Nursing (inc Midwives + HCAs) | 1160 (20.6) | 698 (21.6) | 462 (19.3) | |
| AHPs | 2277 (40.4) | 1301 (40.2) | 976 (40.7) | |
| Dental | 325 (5.8) | 171 (5.3) | 154 (6.4) | |
| Administrative/estates/other | 321 (5.7) | 184 (5.7) | 137 (5.7) | |
| Missing | 185 (3.3) | 103 (3.2) | 82 (3.4) | |

* Responders and non-responders compared using chi-squared tests for categorical variables and Wilcoxon rank-sum tests for continuous variables. Values expressed as n(%) unless stated otherwise.
